# Generative AI avatar videos for tobacco prevention on social media: a randomized controlled trial

**DOI:** 10.64898/2026.06.22.26356232

**Authors:** Nicolas B. Merl, Christoph Wies, Franziska Schramm, Jana T. Winterstein, Tirtha Chanda, Titus J. Brinker

## Abstract

Short-form video platforms increasingly shape how young audiences encounter health information. Generative artificial intelligence can produce standardized avatar-based messages at scale, but randomized evidence for tobacco prevention is scarce. In this three-arm randomized online intervention study with pre-post assessment, participants aged 16 years or older were assigned to an AI avatar video emphasizing short-term smoking consequences, an AI avatar video presenting long-term cancer-related information matched to an American Cancer Society fact sheet, or the same fact sheet in written form. The primary outcome was post-intervention intention to avoid smoking and secondhand smoke exposure, adjusted for baseline intention. Among 400 randomized participants, 272 had complete data for the primary baseline-adjusted analysis. Intention increased from baseline to post-intervention in all conditions, with no statistically significant between-group differences. These findings support AI avatar videos as a scalable, social-media-compatible format for digital tobacco prevention, while not establishing superiority or equivalence.

## Introduction

Tobacco use remains the leading preventable cause of cancer and cancer mortality worldwide, with initiation frequently occurring during adolescence and young adulthood^1^. At the same time, health information consumption has shifted increasingly toward social media platforms dominated by short-form video content, such as Instagram and TikTok^2^. Despite this transition, many established tobacco prevention resources still rely on traditional text-based materials, including fact sheets, brochures, posters, and flyers^3^.

Because communication effectiveness depends not only on message content but also on delivery modality^4^. This is particularly relevant for young social media users, whose everyday content environments are strongly shaped by entertainment- and leisure-oriented formats^5^. It therefore remains unclear whether prevention messages presented in social-media-native video formats are as effective as conventional written materials in influencing tobacco-related attitudes and intentions among young people.

Recent advances in Generative Artificial Intelligence (GenAI) have enabled the scalable production of realistic avatars capable of delivering standardized health communication messages^6^. Such approaches may offer advantages for public health communication, including low-cost content generation, rapid scalability, and adaptation to social-media-specific communication formats. Although emerging evidence suggests that social media content generated by Artificial Intelligence (AI) can increase user engagement^7^, rigorous evidence in tobacco prevention remains limited. Moreover, studies that vary format and content simultaneously make it difficult to determine whether observed effects are attributable to the delivery format itself or to differences in message framing^8^.

This distinction may be particularly relevant in tobacco prevention. While prevention campaigns frequently focus on long-term health risks such as cancer, these outcomes may appear psychologically distant and less personally relevant to younger individuals^9^. By contrast, short-term consequences of smoking, including impacts on appearance, social interactions, physical performance, and financial costs, are more immediate and easier to relate to^10^. As a result, framing smoking consequences in short-term terms may enhance message relevance, engagement, and persuasive impact.

To assess the psychological effects of prevention messaging, this study draws on the Theory of Planned Behaviour (TPB), a widely used framework for explaining health-related intentions^11^. In the TPB, intention is considered the proximal determinant of behaviour, and meta-analytic evidence shows robust associations between intentions and subsequent behavior^12^. According to the TPB, behavioural intentions are shaped by attitudes, perceived social norms, and perceived behavioural control^11^. Communication modality and message framing may influence smoking avoidance intentions by affecting the perceived relevance and persuasiveness of prevention content.

Beyond individual-level persuasion, communication modality may also affect the public health impact of prevention messages. On social media platforms such as Instagram and TikTok, short-form videos are embedded in algorithmically curated feeds and are central to content discovery and engagement on contemporary social media platforms^13^. As younger audiences increasingly obtain health information through these platforms, video-based prevention messages may offer dissemination advantages beyond their direct effects on smoking-related intentions. Furthermore, in digital environments characterized by intense competition for user attention, intervention exposure time is relevant as a process indicator of how users engage with different communication formats^14^. Therefore, intervention exposure time was assessed as an indicator of communication efficiency and feasibility.

We conducted a randomized controlled trial to compare AI avatar-based tobacco prevention videos with a conventional written tobacco prevention fact sheet. One avatar condition was content matched to the fact sheet to isolate communication modality effects, whereas a second emphasized short-term, socially salient smoking consequences. We examined intervention exposure time and dropout as process outcomes to assess whether avatar-based delivery showed different exposure and completion patterns compared with fact sheet-based information, as well as smoking avoidance intentions, TPB-related determinants, perceived knowledge gain, and engagement intentions. We hypothesized that AI avatar-based videos would be associated with immediate changes in smoking and secondhand smoke avoidance intentions and TPB-related determinants, and that these effects would be comparable to those observed in the fact sheet condition.

## Results

### Study design

This study employed a three-arm randomized controlled intervention study with a pre-post assessment design implemented via the platform LimeSurvey^15^. Participants completed a baseline questionnaire assessing TPB constructs related to smoking, were assigned and exposed to one of three intervention conditions, and subsequently completed the same questionnaire immediately post-intervention. The three conditions were: (1) an AI avatar delivering short-term consequences of exposure to tobacco smoke (avatar-short-term), (2) an AI avatar delivering factual framed cancer prevention information that was content-matched to the American Cancer Society (ACS) “Tobacco and Cancer Fact Sheet for Patients and Caregivers”^16^ (avatar-long-term), and (3) the ACS flyer presented in written form as the standard information control (control). Participants in all conditions were required to remain on the intervention page for a minimum exposure time of 45 seconds before proceeding to the post-intervention questionnaire. This interval was chosen to slightly exceed the 40-second avatar video duration, allowing sufficient time for participants to start the video and proceed to the next survey page after viewing the material. After the exposure period, participants could continue to view the assigned material for an unrestricted duration. Participants could continue engaging with the assigned material by reading or replaying it. The primary endpoint was the participants’ intention not to smoke and to avoid exposure to tobacco smoke in the next four weeks at post-test, adjusted for baseline intention derived from pre-test. The study design is shown in figure 1.

**Figure 1.**
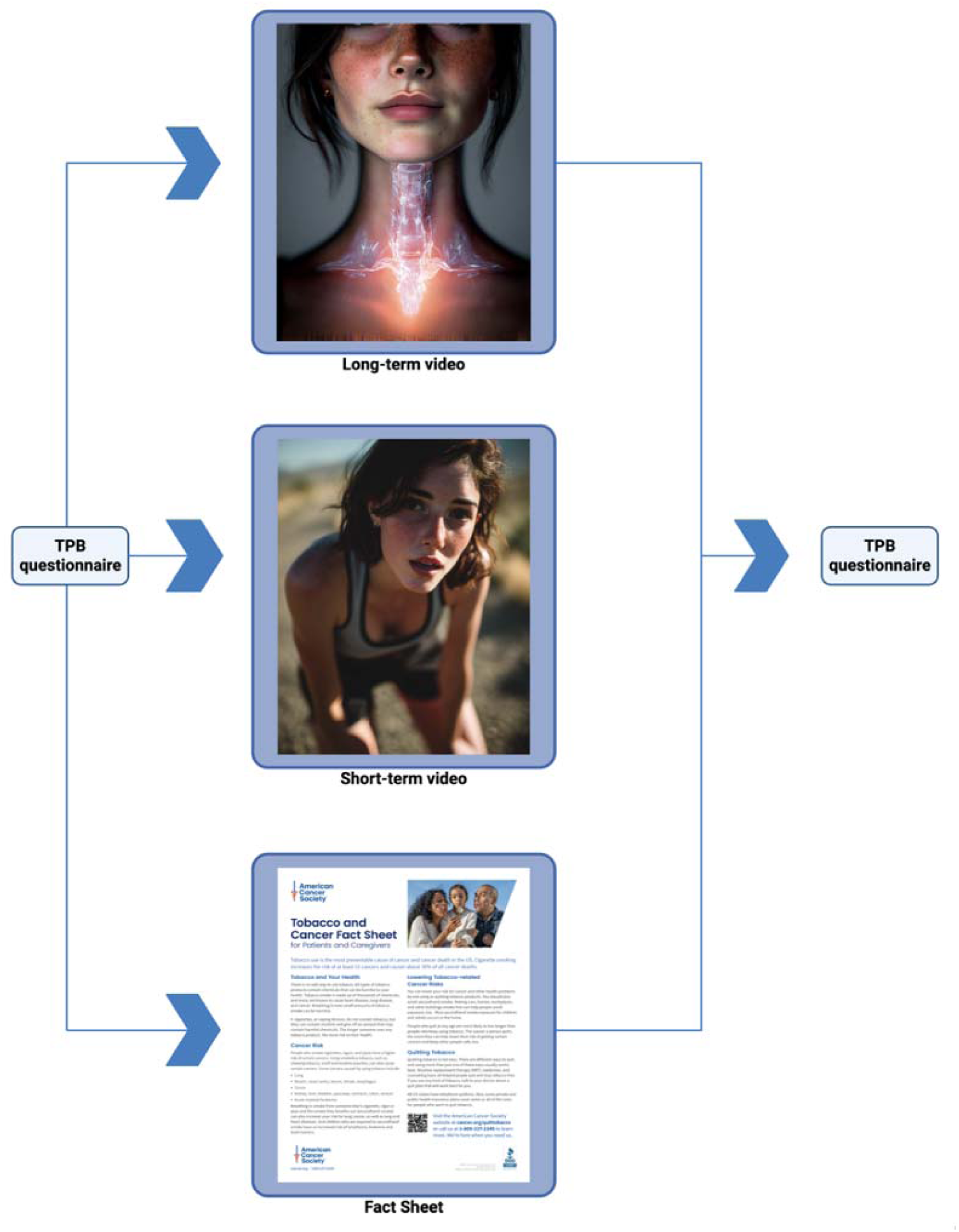
Study design and intervention flow. Participants completed a baseline Theory of Planned Behaviour (TPB) questionnaire and were then randomly assigned to one of three intervention conditions: an AI avatar video presenting long-term tobacco-related cancer risks, an AI avatar video presenting short-term consequences of tobacco use, or a written tobacco and cancer fact sheet. After exposure to the assigned intervention, participants completed the post-intervention TPB questionnaire.

Participant recruitment and data collection were conducted between February 13, 2026, and June 5, 2026. The final analytic sample for the pre-post analyses included 309 participants. Most participants were aged 16–29 years (82.2%), and the sample was slightly more likely to identify as female (53.7%) than male (34.3%). Baseline Theory of Planned Behavior scores were generally high across intervention conditions, with similar distributions for intention, attitude, injunctive norm, and perceived behavioral control. Baseline characteristics and descriptive statistics by intervention condition are shown in Table 1. Figure 2 shows the CONSORT-flow-diagram.

**Figure 2.**
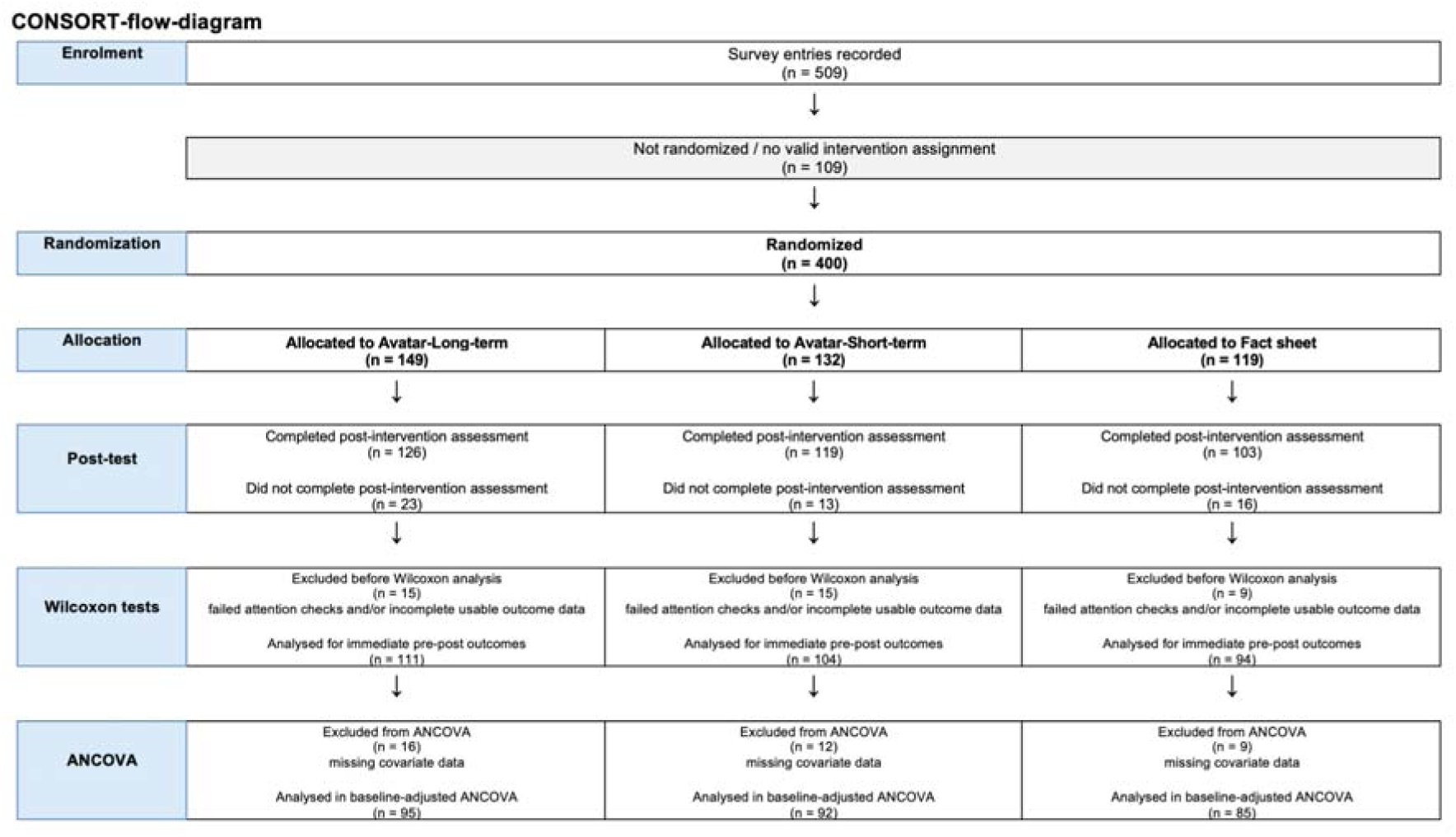
A total of 509 survey entries were recorded. Of these, 400 participants reached randomization and were assigned to one of the three intervention conditions: 149 to avatar-long-term, 132 to avatar-short-term, and 119 to the fact sheet control condition. The final analytic sample for the Wilcoxon signed-rank tests consisted of 309 participants after exclusion of incomplete responses and participants who failed both attention checks. (avatar-Long-term n = 111, avatar-Short-term n = 104, fact sheet control n = 94). The baseline-adjusted ANCOVA models additionally required complete covariate data for age group, gender, smoking status, and education; due to missing covariate values, these models were based on n = 272 participants (avatar-Long-term n = 95, avatar-Short-term n = 92, fact sheet control n = 85). The final sample size for ANCOVA is smaller than planned due to the unexpectedly high number of participants who withdrew.

**Table 1.**
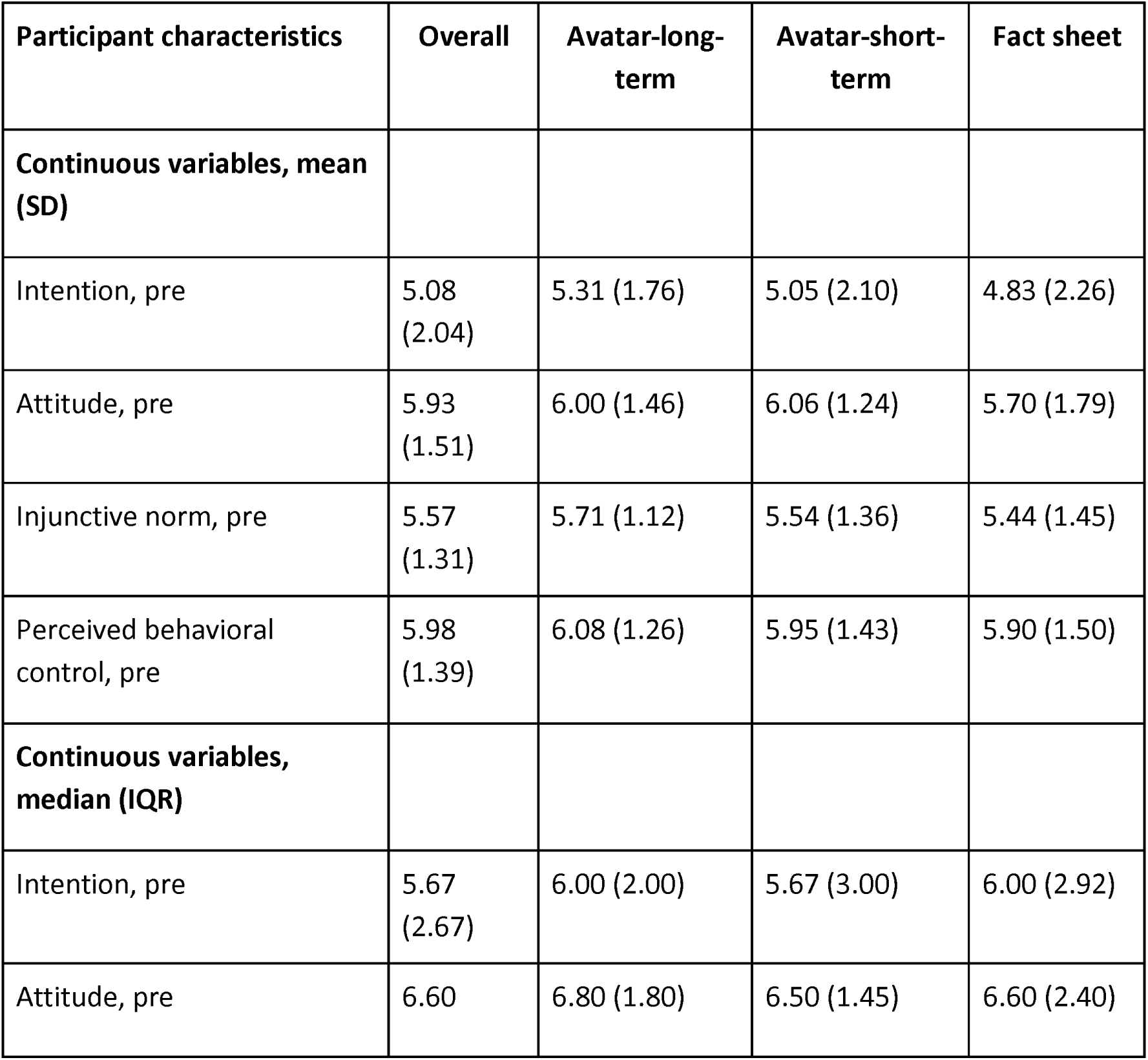
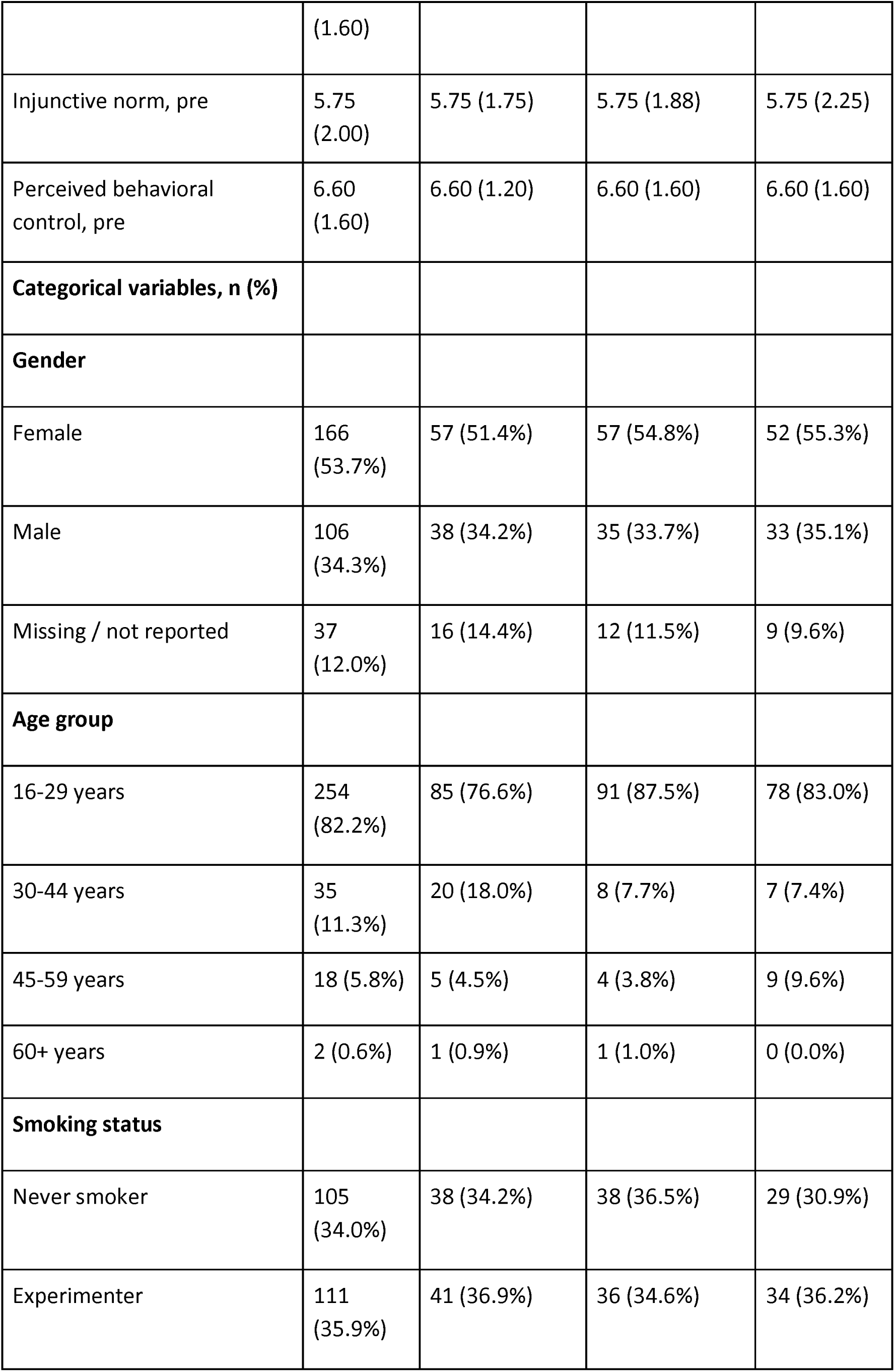
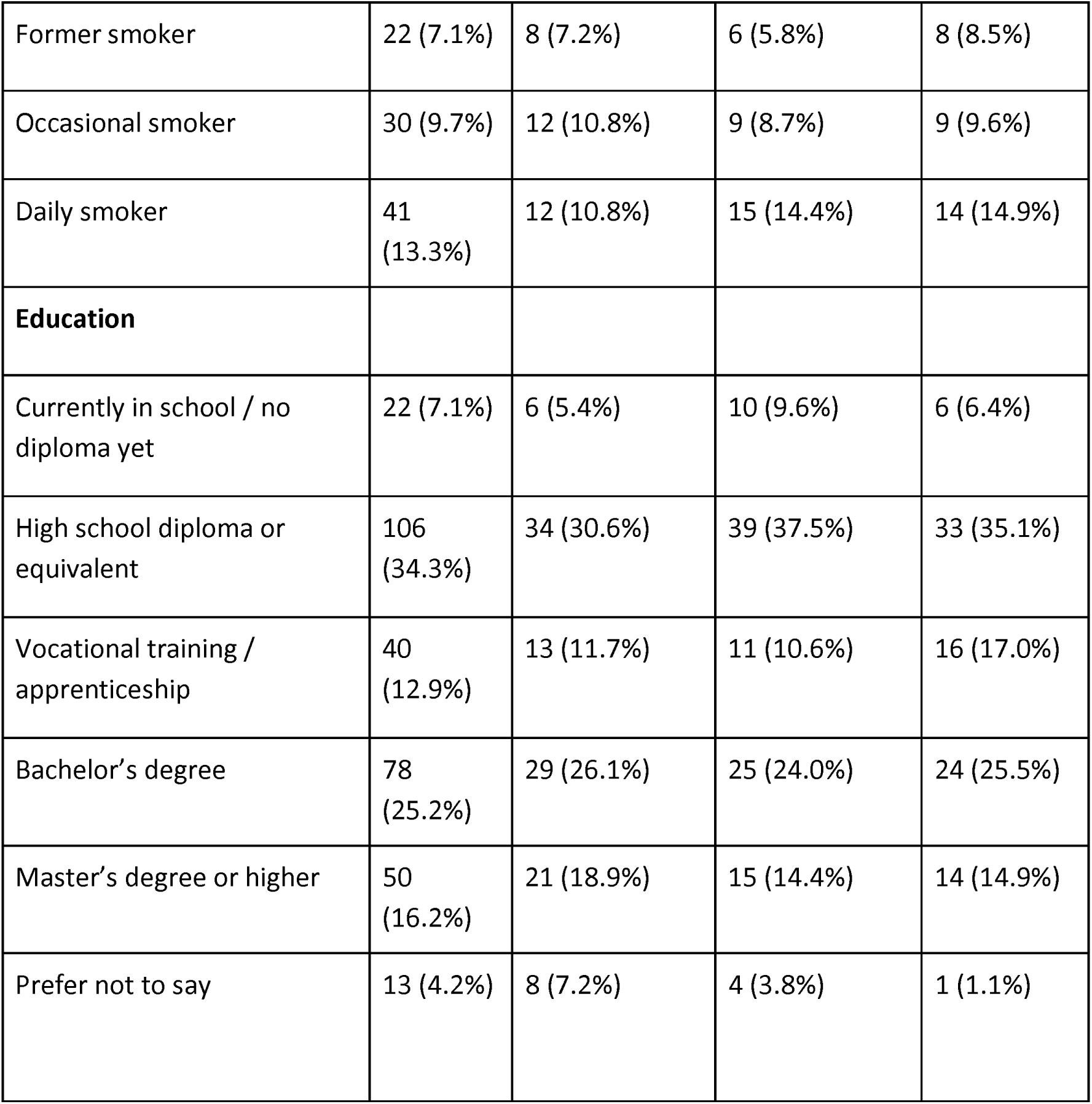
Baseline characteristics and descriptive statistics by intervention condition. Values are n (%) unless otherwise indicated. Baseline Theory of Planned Behavior scale scores are reported as both mean (SD) and median (IQR). Percentages are calculated within each intervention condition. The table describes the final analytic sample for pre-post analyses, n = 309.

Internal consistency was good to excellent for intention, attitude, and perceived behavioural control at both measurement points. Cronbach’s alpha (α) ranged from α = 0.839 to α = 0.913 for these scales. In contrast, the injunctive norm scale showed low internal consistency at baseline, α = 0.553, and post-intervention, α = 0.511; findings for this outcome should therefore be interpreted cautiously.

Manipulation-check ratings were generally high across conditions, supporting the validity of the experimental manipulation and indicating that participants recognized the intended message content. This strengthens the internal validity of the comparisons. Mean (SD) ratings were 5.04 (1.97) in avatar-long-term, 5.80 (1.56) in avatar-short-term, and 5.76 (1.57) in the fact sheet control condition.

Wilcoxon signed-rank tests were used to examine pre-post changes within each intervention condition. Intention to avoid smoking and secondhand smoke exposure increased significantly in all three groups: avatar-long-term, p < 0.001, r = 0.463; avatar-short-term, p < 0.001, r = 0.349; and fact sheet control, p < 0.001, r = 0.480. These within-condition changes indicate short-term improvements after exposure to all prevention materials. The results are presented in Table 2 and graphically illustrated in Figure 3.

**Figure 3.**
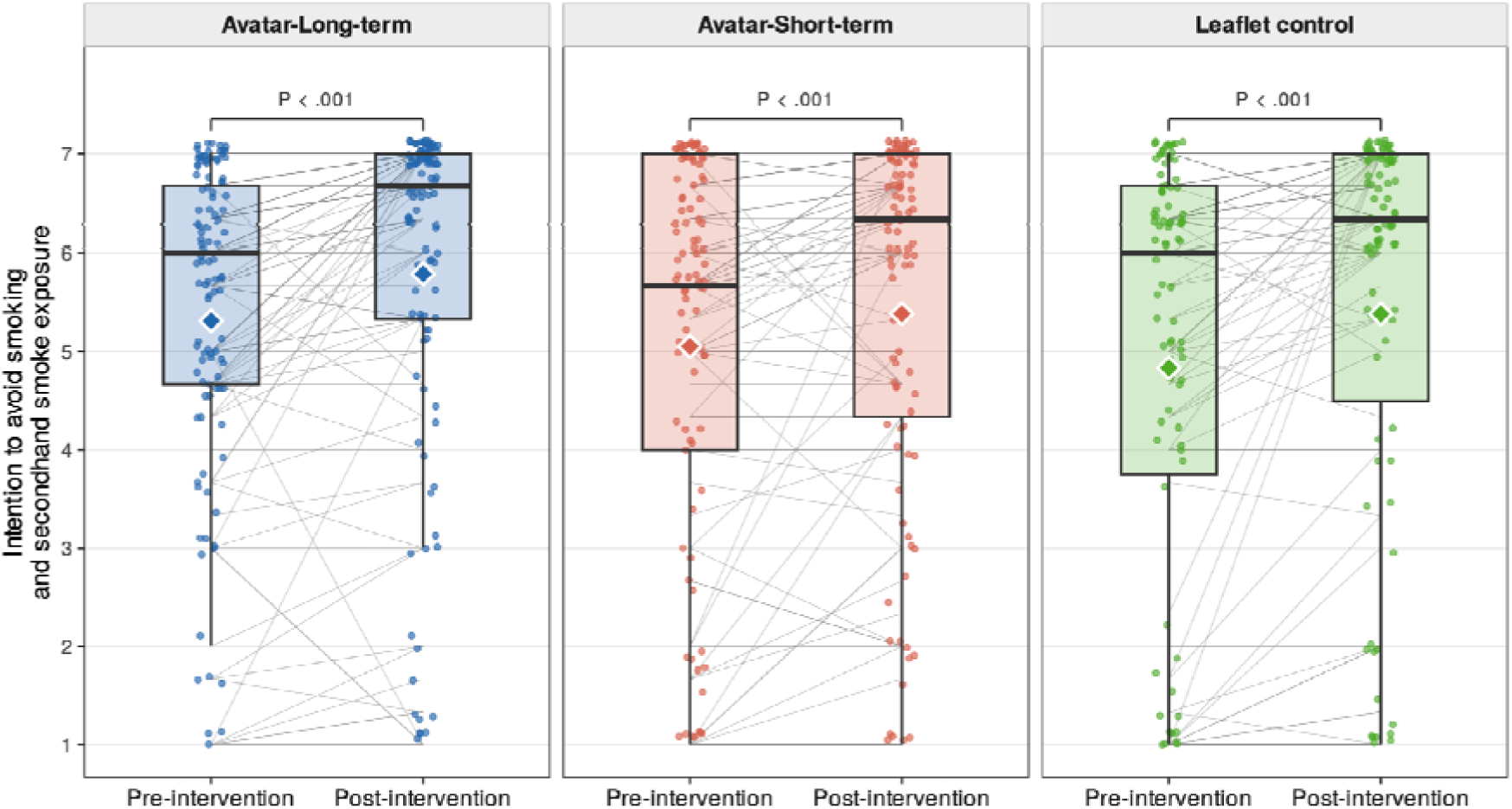
Each panel shows scores for one intervention condition (avatar-Long-term n = 111, avatar-Short-term n = 104, fact sheet control n = 94). Boxes indicate the interquartile range (IQR); the horizontal line within each box indicates the median; whiskers extend to 1.5 × IQR. Diamond symbols indicate individual group means. Grey lines connect pre- and post-intervention scores of the same participant.

**Table 2.**
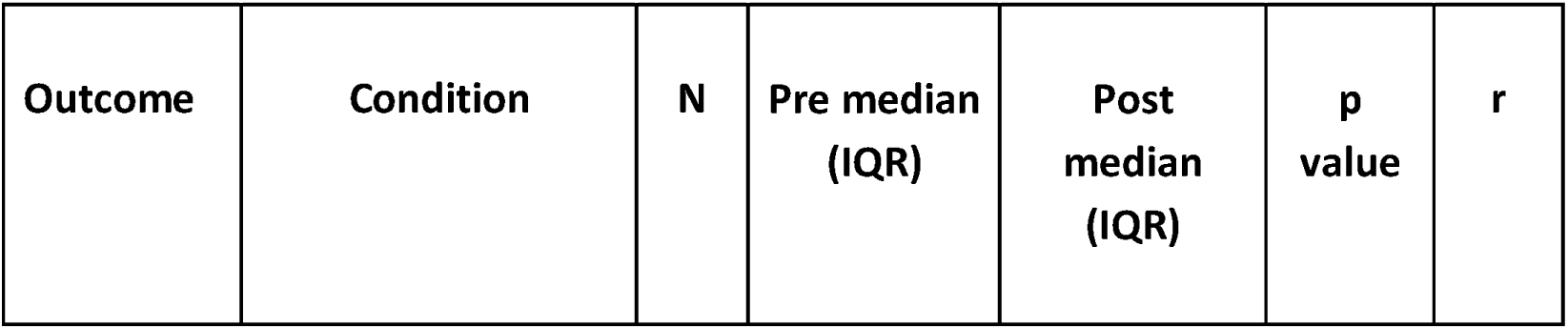
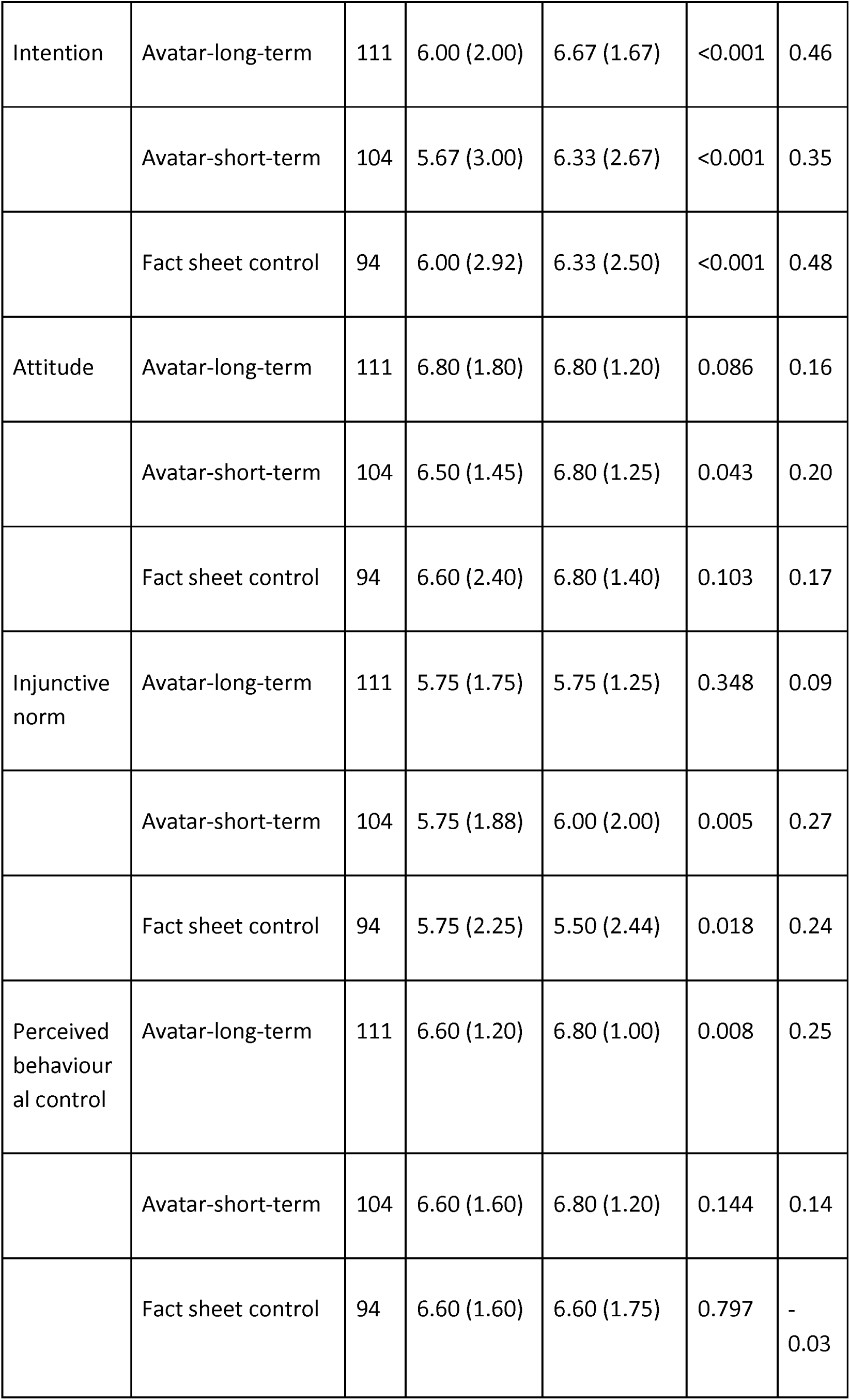
Within-condition pre-post changes in primary and secondary outcomes. Values are medians with interquartile ranges (IQRs). Within-condition changes from baseline to post-intervention were tested using Wilcoxon signed-rank tests. Effect sizes are reported as r, with positive values indicating higher post-intervention scores compared with baseline.

The primary between-condition analyses used baseline-adjusted ANCOVA models including intervention condition, the respective baseline outcome, gender, age group, smoking status, and education. Due to missing covariate data, the ANCOVA models were based on n = 272 participants. ANCOVA results are presented in Table 3.

**Table 3.**
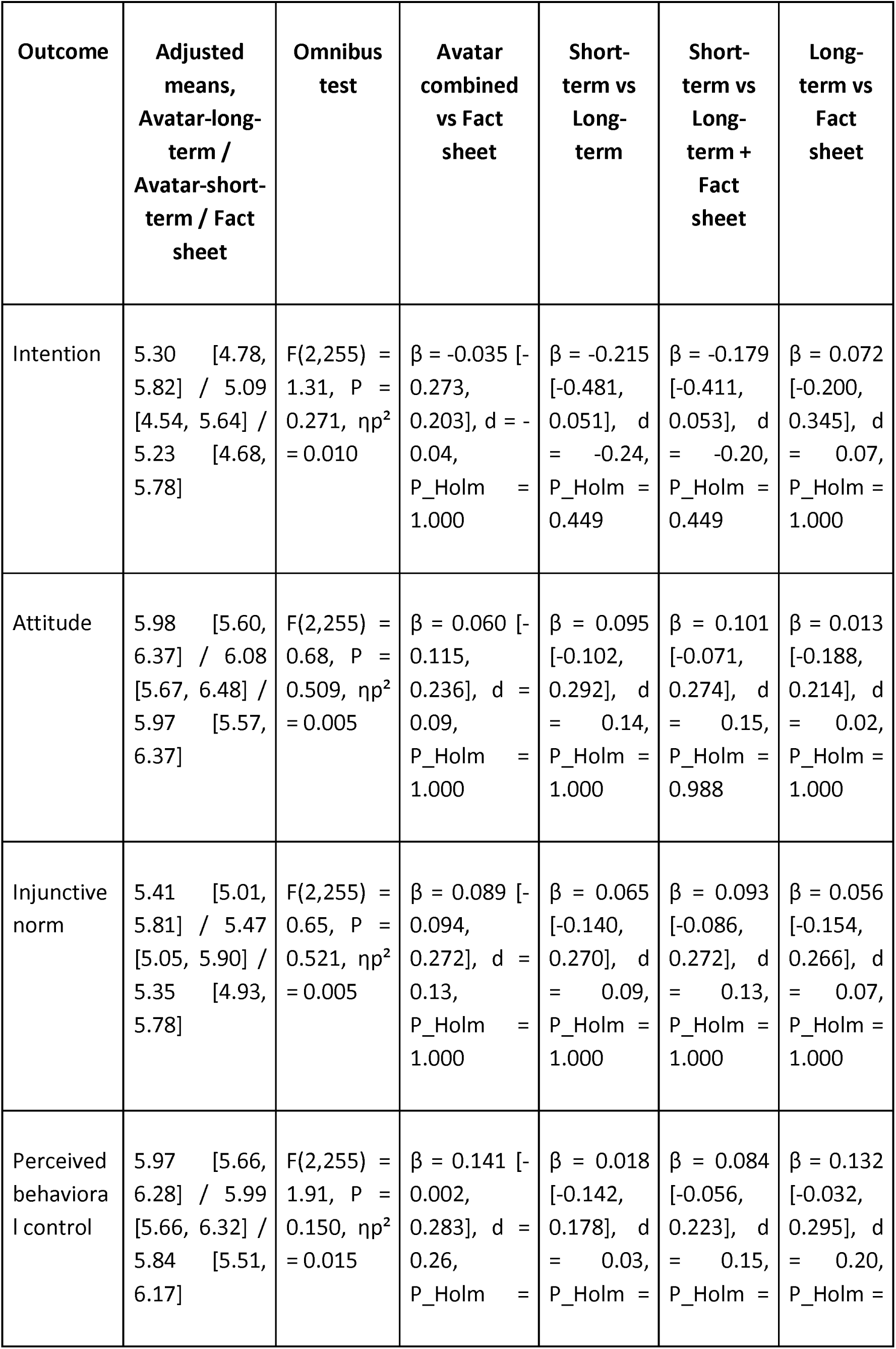
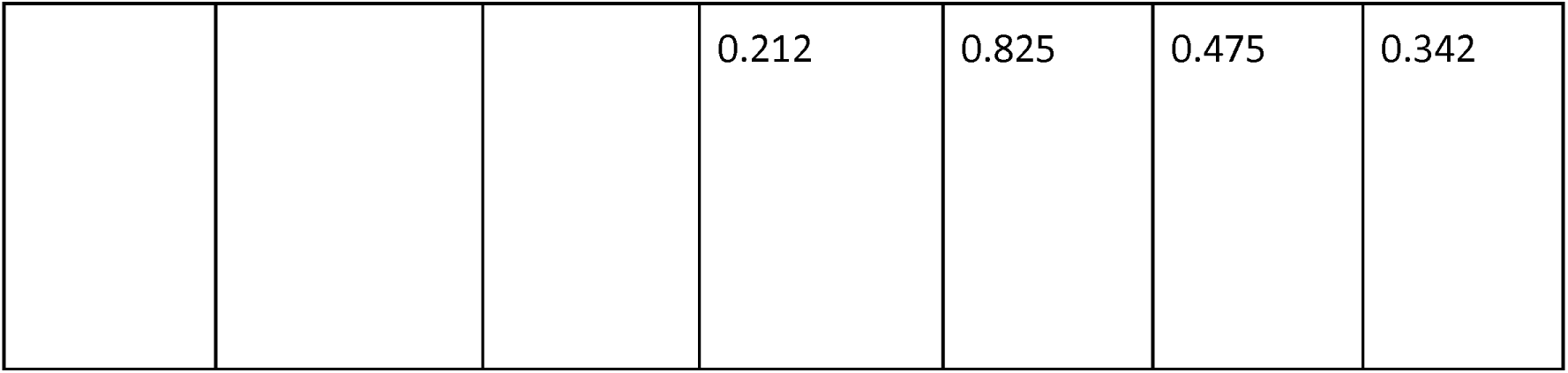
Baseline-adjusted ANCOVA results and planned contrasts for post-intervention outcomes. Values are based on baseline-adjusted ANCOVA models including intervention condition, the respective baseline outcome score, categorical age group, gender, smoking status and education as covariates. Adjusted means are estimated marginal means with 95% confidence intervals. The ANCOVA models were based on n = 272 complete cases. β indicates the adjusted mean difference for each contrast. The four prespecified planned contrasts compared: (1) the two avatar conditions combined versus the fact sheet control condition, (2) avatar-short-term versus avatar-long-term, (3) avatar-short-term versus the avatar-long-term and fact sheet control and (4) avatar-long-term versus fact sheet control contrast. P values for these prespecified contrasts were Holm-adjusted within each outcome. Cohen’s d indicates the standardized effect size for each contrast; corresponding 95% confidence intervals are reported where available. ηp² indicates partial η² for the omnibus condition effect. No omnibus intervention effect or planned contrast reached statistical significance.

For the primary outcome, intention, the omnibus effect of intervention condition was not significant, p = 0.271, partial η² = 0.010. There were also no significant condition effects for attitude, p = 0.509, partial η² = 0.005; injunctive norm, p = 0.521, partial η² = 0.005; or perceived behavioural control, p = 0.150, partial η² = 0.015.

Planned contrasts based on estimated marginal means showed no significant differences for the primary outcome. The two avatar conditions combined did not differ from the fact sheet control condition (estimate = -0.035; 95% CI: -0.273 to 0.203; p_Holm = 0.772). Avatar-short-term did not differ from avatar-long-term (estimate = -0.215; 95% CI: -0.481 to 0.051; p_Holm = 0.337). Avatar-short-term did not differ from the average of avatar-long-term and fact sheet control (estimate = -0.179; 95% CI: -0.411 to 0.053; p_Holm = 0.337). Avatar-long-term did not differ from fact sheet control (estimate = 0.073,95%; CI: -0.2 to 0.345; p_Holm = 1.0).

Planned contrasts for the secondary outcomes were also non-significant after Holm correction. The closest result was observed for perceived behavioural control, where the combined avatar conditions showed descriptively higher adjusted scores than fact sheet control, estimate = 0.141, p = .053, pHolm = 0.159.

Intervention exposure time was available for 371 of the 400 randomized participants. Mean exposure time was 71.90 seconds in the avatar-long-term condition, 69.11 seconds in the avatar-short-term condition, and 95.81 seconds in the fact sheet control condition, while median exposure times were similar across conditions. The log-transformed exposure-time model showed no significant effect of intervention condition (p = 0.193, partial η² = 0.009).

Overall, 52 of 400 randomized participants were classified as dropouts, corresponding to a dropout rate of 13.0%. Dropout rates were 15.4% in avatar-long-term, 9.8% in avatar-short-term, and 13.4% in fact sheet control. Logistic regression showed no significant overall effect of condition on dropout, P = 0.365. Compared with avatar-long-term, the odds of dropout were descriptively lower in avatar-short-term, OR = 0.60, 95% CI [0.29, 1.24], P = 0.165, and similar in fact sheet control, OR = 0.85, 95% CI [0.43, 1.70], P = 0.646. Planned dropout contrasts were also non-significant after Holm correction. The contrast comparing avatar-short-term with the two more cancer-focused conditions was not significant, estimate = -0.046, 95% CI [-0.112, 0.020], P = 0.173.

Model diagnostics did not indicate major violations of ANCOVA assumptions for the primary outcome. The homogeneity of regression slopes assumption was supported for intention, injunctive norm, and perceived behavioural control, whereas the condition × baseline interaction was significant for attitude.

For attitude, the condition × baseline interaction was significant, indicating a potential violation of the homogeneity of regression slopes assumption. Consequently, the attitude results are reported descriptively and should be interpreted with caution. This did not affect the primary outcome analysis, for which the assumption was supported.

Sensitivity analyses using baseline-adjusted ANCOVA models without demographic covariates yielded the same pattern of findings, with no significant omnibus intervention effect for intention (p = 0.334), attitude (p = 0.894), injunctive norm (p = 0.553), or perceived behavioural control (p = 0.152). Thus, results were robust to excluding demographic covariates.

Overall, intention to avoid smoking and secondhand smoke exposure increased significantly within all three conditions. However, the baseline-adjusted between-condition analyses and planned contrasts showed no significant differences between the AI avatar videos and the fact sheet control, nor between short-term and long-term avatar framing. Exposure time and dropout also did not differ significantly between conditions.

## Discussion

This randomized controlled trial examined whether GenAI avatar videos in a social-media-native format can be used for tobacco prevention. The findings indicate that AI-generated avatar videos produced short-term changes in smoking-related prevention outcomes that were comparable to those observed after exposure to an established fact sheet-based control condition. Intention to avoid smoking and secondhand smoke exposure increased significantly from pre- to post-intervention in all three conditions. Baseline-adjusted between-group analyses did not show statistically significant differences between the avatar conditions and the fact sheet control, nor between short-term and long-term avatar framing.

These results are relevant for digital public health because they suggest that evidence-based tobacco prevention messages can be translated into AI-generated, short-form avatar videos without loss of immediate intentional impact compared with conventional written material. This is important because young people increasingly encounter health information in visually driven, platform-native environments such as Instagram, TikTok, and other short-form video channels^17^. Even when individual-level effects are comparable to those of written information, avatar videos may offer advantages for implementation, including scalability, standardization, rapid adaptation, and compatibility with social media dissemination^18^. For public health campaigns, this creates an opportunity to disseminate prevention messages in formats that may be more visible, shareable, and acceptable in social media settings^7^.

The comparison between the avatar-short-term and avatar-long-term conditions did not show a significant advantage of short-term framing. Both avatar formats performed similarly on the primary and secondary outcomes. This suggests that AI avatar videos may be suitable for delivering both immediate consequence framing and long-term cancer prevention information. Future studies should examine whether framing effects become stronger under real-world social media conditions, repeated exposure, or with outcomes such as retention, sharing, engagement, and behavioural follow-up.

Secondary outcomes supported the feasibility of avatar-based delivery under controlled study conditions. Although participants were required to remain on the intervention page for a minimum period, median exposure times exceeded this threshold across all conditions, suggesting that participants generally engaged with the assigned material beyond the required exposure period regardless of the intervention received. Dropout did not differ significantly between conditions, providing no evidence that avatar-based delivery was associated with lower completion or reduced exposure compared with ACS leaflet-based information. In digital prevention, such implementation-related outcomes are important because scalable interventions must be not only persuasive but also acceptable and usable^19^.

Intention to avoid smoking and secondhand smoke exposure increased across all conditions, whereas attitude, injunctive norms, and perceived behavioural control did not show consistent changes. This may indicate that brief tobacco prevention messages can trigger immediate motivational responses without necessarily changing broader underlying TPB determinants. These constructs may be more stable and may require repeated exposure, stronger personalization, or more interactive intervention components to shift.

Several limitations should be considered. First, the outcomes were psychological determinants of short-term smoking avoidance rather than long-term behavioural endpoints. At the same time, social-media-based interventions may be particularly suited for repeated exposure and large-scale dissemination. Second, the final analytic models were based on complete cases with available covariate data, which reduced the sample size for ANCOVA analyses. Third, the injunctive norm scale showed low internal consistency and should be interpreted cautiously. As a result, findings for injunctive norms should be interpreted cautiously and should not be considered strong evidence for or against intervention effects on perceived social norms. Finally, although the ANCOVA analyses included marginally fewer complete cases than originally targeted due to missing covariate data, the observed between-condition effects were small and consistently non-significant, supporting the conclusion that neither avatar condition showed a clear advantage over the leaflet control.

Overall, the findings support the potential of GenAI avatar videos as a scalable format for tobacco prevention in social media environments, while also underscoring that comparable immediate effects should not be interpreted as superiority over established written information. This study contributes to digital public health by showing that evidence-based prevention content can be translated into a short-form, platform-native avatar format without statistically detectable loss of immediate intentional impact in this sample. Such formats may offer implementation advantages, including standardization, rapid adaptation, subtitling, and social media dissemination. Future studies should test these interventions under real-world platform conditions and assess algorithmic reach, sharing, repeated exposure, longer-term behavioural outcomes, and equity effects to determine their public health value beyond controlled experimental settings.

## Methods

### Trial registration and reporting guidelines

The study was registered on the Open Science Framework after data collection but before confirmatory data analysis. The registration specified the study design, intervention conditions, hypotheses, primary and secondary outcomes, and planned statistical analyses at https://osf.io/egbvc/.

Ethics approval for this study was obtained from the Ethics Committee of the Medical Faculty of Heidelberg University (Universität Heidelberg, Ethikkommission der Med. Fakultät), approval number S-626/2025.

This manuscript was prepared in accordance with the CONSORT 2025 reporting guideline for randomized trials. A completed CONSORT-flow chart (Appendix 1) is provided as supplementary material.

### Participants and recruitment

Participants were recruited via Instagram ads and from German vocational schools and Universities to obtain a sample of young people, as broad as possible. Eligibility criteria were age ≥16 years. Participants younger than 16 years were not included because the study was conducted as an anonymous online survey. In this setting, obtaining and verifying parental consent was not possible. No additional eligibility criteria were applied to include a broad range of participants and to approximate the heterogeneity of audiences typically reached in real-world social media environments.

Participants were randomly allocated in a 1:1:1 ratio to one of the three conditions using a computer-generated randomisation procedure implemented in LimeSurvey. Allocation was revealed only after completion of baseline measures. Due to the nature of the interventions, participant blinding was not feasible. Outcomes were self-administered via standardized questionnaires and were identical across conditions.

### Interventions

#### ACS fact sheet control condition (written format)

Participants received the ACS “Tobacco and Cancer Fact Sheet for Patients and Caregivers” in digital written form (single-page fact sheet) (Appendix 2), providing evidence-based smoking prevention information. Participants were instructed to read the fact sheet carefully. Exposure time was standardised by a minimum time-on-page requirement of 45 seconds (matching the time from the other two conditions), after which participants could continue to the post-test questionnaire.

#### Avatar-long-term condition (ACS content-matched, spoken format)

Participants viewed an AI avatar delivering a brief factual cancer prevention message. The avatar script (Appendix 3) was systematically derived from and closely aligned with the fact sheet, with the explicit aim of matching the fact sheet’s informational content while differing only in delivery format (spoken avatar versus written text). Core informational components reflected the fact sheet, including (i) the statement that tobacco use is a leading preventable cause of cancer and cancer death, (ii) that there is no safe way to use tobacco and it contains thousands of harmful chemicals, (iii) that smoking increases the risk of multiple cancers and that smokeless tobacco also causes cancer, and (iv) that secondhand smoke increases health risks, including cancer risk. Participants were instructed to watch the avatar video carefully. The duration of the avatar message was matched to the planned exposure time of the fact sheet condition.

#### Avatar-short-term condition (direct consequences of smoking, spoken format)

Participants viewed the same AI avatar as in the avatar-long-term intervention delivering a brief cancer prevention message framed around direct short-term consequences of smoking (Appendix 4). In contrast to the avatar-long-term condition, this script focused on short-term consequences of smoking containing (i) social norms: smoking is associated with perceived social disapproval and social distancing by peers, (ii) financial costs: smoking is associated with substantial cumulative financial costs, potentially reducing disposable income for other needs or activities, (iii) rapid appearance and sensory consequences: smoking can contribute to unpleasant odour, tooth discoloration, and acute symptoms such as headaches, and (iv) reduced performance in sports and everyday life: smoking is associated with impaired cardiorespiratory fitness, which can manifest as breathlessness during exercise and daily activities. The duration and visual presentation were matched to the avatar-long-term condition and the planned exposure time of the fact sheet condition.

### Standardization across conditions

The two avatar conditions used the same avatar appearance and presentation style and were matched in message duration (40s). The avatar-long-term condition was explicitly content matched to the fact sheet to allow a direct comparison of delivery modality under equivalent informational content. For example, the information that secondhand smoke is harmful was included in the written brochure and was also mentioned in the avatar-long-term video. The avatar-short-term condition differed in message content to test the effect of emphasizing short-term versus long-term smoking consequences within the avatar modality.

Both avatar based interventions were delivered as videos designed to resemble short-form social media content, such as Instagram Reels and YouTube Shorts. The avatar videos were produced in a 4:5 vertical format suitable for mobile viewing and commonly used in social media contexts. Participation was possible on both smartphones and computers. Participants were instructed to enable audio during video playback to ensure full exposure to the auditory content. In addition, on screen subtitles were presented to enhance comprehension in muted playback contexts and to ensure that key message content was conveyed regardless of audio use. The visual layout and pacing were standardized across the avatar short-term and avatar long-term conditions, including the same avatar appearance, background, video length, and overall editing style, to ensure that the primary differences between avatar conditions were limited to message framing and content.

### Measures

#### Theory of Planned Behaviour

TPB constructs were assessed using a questionnaire (Appendix 5) developed in accordance with established TPB measurement guidance and tailored to the target behaviour (“not smoking and avoiding smoke exposure”) within a fixed four-week time window. All Likert-type items used a 7-point response scale (1 = strongly disagree to 7 = strongly agree). Attitudes were measured via semantic differential items on 7-point bipolar scales.

Intention to avoid smoking and secondhand smoke exposure in the next four weeks was assessed with three items (1.1 - 1.3). Mean scores were computed, with higher values indicating stronger non-smoking and passive smoking avoidance intention. Attitude toward avoiding smoking and secondhand smoke exposure was assessed via five semantic differential items (2.1 - 2.5), averaged to form an attitude score. Injunctive norm was assessed with four items (3.1 - 3.4) capturing perceived expectations, approval, and disapproval by important others. The disapproval item was reverse-coded so that higher values reflect stronger injunctive norm for non-smoking. Mean scores were computed. Perceived behavioural control (PBC) was assessed with five items (4.1 - 4.5) reflecting perceived ease, self-efficacy, and controllability of not smoking, averaged to form a PBC score.

Internal consistency was assessed using Cronbach’s alpha (separately for baseline and post-test).

#### Intervention exposure time and after intervention dropouts

LimeSurvey automatically recorded the duration participants remained on the intervention page before advancing to the post-intervention questionnaire. Exposure time was used as a process indicator of engagement with the intervention material and was compared across study conditions. Participants were classified as dropouts if they had been exposed to one of the intervention conditions but did not complete the study through the final post-intervention questionnaire.

To verify that participants perceived the intended framing of the intervention, post-intervention manipulation checks assessed perceived message emphasis using condition-specific items. Participants in the avatar-short-term condition received the item: “The message mainly focused on short-term consequences of smoking, such as social expectations, financial costs, physical performance, and sensory consequences.” Participants in the avatar-long-term and fact sheet control conditions received the item: “The message mainly focused on smoking-related cancer risk.” Each item was rated on a 7-point Likert scale.

#### Sociodemographic and smoking-related variables

After the questionnaire, participants reported age group, gender and highest education degree. Smoking status was assessed at baseline. The reported age-group was categorized as 16-29, 30-44, 45-59, 60+ years and, smoking status was modelled as never smoker, experimenter, former smoker, occasional smoker, daily smoker., Education was assessed as “Currently in school / no diploma yet”, “High school diploma or equivalent”, “Vocational training / apprenticeship”, “Bachelor’s degree”, “Master’s degree or higher”, or “Prefer not to say”.

#### Data quality and exclusions and Manipulation checks

Quality control procedures were prespecified. The survey included two attention check items. Participants were excluded from primary analyses if they failed both attention checks. Additionally, participants with missing TPB outcome data post-test were excluded for primary TPB related outcomes.

To verify that participants perceived the intended framing of the intervention, post-intervention manipulation checks assessed perceived message emphasis using condition-specific items. Participants in the avatar-short-term condition received the item: “The message mainly focused on short-term consequences of smoking, such as social expectations, financial costs, physical performance, and sensory consequences.” Participants in the avatar-long-term and fact sheet control conditions received the item: “The message mainly focused on smoking-related cancer risk.” Each item was rated on a 7-point Likert scale.

#### Outcomes

The primary outcome was the participants’ post-intervention intention to avoid smoking and secondhand smoke exposure in the next four weeks, adjusted for baseline intentions. Secondary outcomes were post-intervention attitude, injunctive norm, and PBC, each adjusted for baseline values. Additional secondary outcomes included the exposure time and dropout rate after intervention.

#### Sample size

A priori sample size planning targeted adequate power to detect a small-to-moderate between-group effect in the baseline-adjusted primary outcome analysis. Because no prior randomized studies were available to provide a precise estimate of the expected effect in this specific context, the sample size calculation was based on a small-to-moderate omnibus effect of Cohen’s f = 0.20 for the three-group ANCOVA, corresponding to ηp² ≈ .038. With α = 0.05 and 80% power, the required sample size was estimated at approximately 100 participants per condition. To account for exclusions due to incomplete responses and data quality criteria, the recruitment target was increased by approximately 20%, resulting in a planned total sample size of approximately 360 participants, corresponding to 120 participants per condition.

### Statistical analysis

All analyses were prespecified and conducted in R (version 4.3.3), including the package emmeans (version 2.0.3). Statistical significance was set at α = 0.05 (two-sided) for the primary outcome. Scale scores were computed as mean item ratings. Internal consistency was assessed using Cronbach’s alpha at baseline and post-test. P-values for all planned contrasts were adjusted using the Holm correction within each outcome. Unless otherwise specified, 95% confidence intervals were calculated using model-based standard errors from the corresponding fitted statistical models. For logistic regression analyses, odds ratios and corresponding 95% confidence intervals were derived from the fitted logistic regression model.

#### Primary analysis

The primary outcome was post-intervention intention to avoid smoking and secondhand smoke exposure over the subsequent four weeks. Within-group changes from baseline to post-intervention were examined separately for each intervention condition using Wilcoxon signed-rank tests. For these within-group tests, effect sizes were reported Wilcoxon effect size r. These analyses described immediate pre-post changes in smoking and secondhand smoke avoidance intentions and TPB determinants after exposure to each prevention format.

Group differences were examined using baseline-adjusted analysis of covariance (ANCOVA), with study condition specified as a fixed factor with three levels. Baseline intention was entered as a covariate. To increase statistical precision and account for baseline heterogeneity, age group, gender, smoking status, and education were additionally included as prespecified covariates.

Adjusted group differences were expressed as estimated marginal means (EMMs) with corresponding model-based 95% confidence intervals (CI). Four a priori planned contrasts were tested:

i. The two avatar conditions combined versus the fact sheet control (delivery modality effect).
ii. The avatar-short-term versus avatar-long-term (framing effect within the avatar modality).
iii. The avatar-long-term and fact sheet vs. avatar-short-term (framing effect between modalities).
iv. The avatar-long-term versus the fact sheet control (delivery modality effect for long-term consequences)

Effect sizes were reported as partial η² for omnibus tests and Cohen’s d for planned contrasts based on adjusted means.

#### Secondary analyses and robustness checks

Secondary TPB outcomes, including attitude, injunctive norm, and perceived behavioural control, were analysed using baseline-adjusted ANCOVA models analogous to the primary outcome analysis. For each outcome, the post-intervention score was entered as the dependent variable, with intervention condition, the corresponding baseline score, age group, gender, smoking status, and education included as covariates. The same planned contrasts were applied to the secondary outcomes.

Intervention exposure time and dropout were analysed as additional process outcomes. Because exposure time was right-skewed, between-group differences were examined using a linear model with log-transformed exposure time as the dependent variable and intervention condition as the predictor. Dropout after randomization was analysed using logistic regression with intervention condition as the predictor. Odds ratios with 95% CIs were reported.

For each ANCOVA model, residuals-versus-fitted plots were inspected for systematic residual patterns, Q-Q plots for approximate normality of residuals, scale-location plots for homogeneity of residual variance, and Cook’s distance plots for influential observations. The homogeneity of regression slopes assumption was assessed by adding and testing a Condition × baseline outcome interaction term in each outcome-specific model.

As a sensitivity analysis, the baseline-adjusted ANCOVA models were repeated without demographic covariates, including only intervention condition and the respective baseline outcome score. This analysis was conducted to assess whether the results were robust to the reduced complete-case sample caused by missing demographic covariate data.

## Statement on the use of AI tools

In accordance with the COPE (Committee on Publication Ethics) position statement of 13 February 2023 (https://publicationethics.org/cope-position-statements/ai-author), the authors hereby disclose the use of the following AI models during the writing of this article: GPT-5.5 (ChatGPT, OpenAI) for checking and improving spelling and grammar.

## Data Availability

All data, analysis code, and study materials are publicly available at: https://zenodo.org/records/20761083?preview=1&token=eyJhbGciOiJIUzUxMiJ9.eyJpZCI6IjE4NTNjMmFjLTkzZjAtNDA4My05NWQ5LWIyNTE1ODQ3OTNkNyIsImRhdGEiOnt9LCJyYW5kb20iOiJlMmMyOGY2N2NiY2EwYTdhYmFjOGEyM2ZkMTUxZWFkYSJ9.kjIQb3j0ZhH7cb8GE1PEeN7wyCBjfGpv5TXTb2v-IYRW91Yc7NLv3DntSdyvcKzf8lvcHBmVyDEZqZ3GaYZcJA

## Author contributions

N.B.M. developed the study concept, designed the methodology, conducted the investigation, performed the formal analysis, validated the findings, authored the original draft, and created visual representations. F.S. and J.T.W. reviewed and edited the manuscript. C.W. and T.C. created visual representations, contributed to the validation process, participated in the review and editing of the manuscript and performed the formal analysis. T.J.B. led the conceptualization, provided resources, reviewed and edited the manuscript, supervised the project team, administered the project, and acquired funding.

## Acknowledgements

This research was funded by the German Heart Association (Deutsche Herzstiftung e.V.), Frankfurt am Main, Germany (grant holder: TJB); the funder had no involvement in the study design, data collection, analysis, interpretation, or the writing of the manuscript.

