## Appendix 1-5 for "Generative AI avatar videos for tobacco prevention on social media: a randomized controlled trial": Appendix 5.docx

**Theory of Planned Behavior Questionnaire**

*Smoking and Secondhand Smoke Avoidance Intentions*

**Target behavior and time frame:** not smoking and avoiding exposure to tobacco smoke over the next four weeks.

**Response format:** Unless otherwise stated, items are rated on a 7-point scale from 1 = strongly disagree to 7 = strongly agree. Higher mean scores indicate stronger endorsement of the respective construct.

**Scoring note:** Items within each construct are averaged. The subjective norm item marked with an asterisk is reverse-coded before scale calculation.

**Intention**

Please indicate how much you agree with the following statements.

| **Item** | **Question / bipolar endpoints** | **Scale** | **Coding** |
| --- | --- | --- | --- |
| I1 | I plan not to smoke in the next four weeks. | 1 = strongly disagree; 7 = strongly agree | Regular |
| I2 | I intend to refuse cigarettes or vapes if someone offers me one in the next four weeks. | 1 = strongly disagree; 7 = strongly agree | Regular |
| I3 | I plan to avoid situations in which I would passively inhale tobacco smoke over the next four weeks. | 1 = strongly disagree; 7 = strongly agree | Regular |

**Attitude**

Please indicate how you evaluate the behavior by choosing a position on the scale between the two opposite endpoints. For me, deliberately avoiding tobacco smoke in the next four weeks (i.e., not smoking myself and avoiding secondhand smoke) would be...

| **Item** | **Question / bipolar endpoints** | **Scale** | **Coding** |
| --- | --- | --- | --- |
| A1 | bad – good | 1 = left endpoint; 7 = right endpoint | Regular |
| A2 | harmful – beneficial | 1 = left endpoint; 7 = right endpoint | Regular |
| A3 | unpleasant – pleasant | 1 = left endpoint; 7 = right endpoint | Regular |
| A4 | useless – useful | 1 = left endpoint; 7 = right endpoint | Regular |
| A5 | restrictive – unrestrictive | 1 = left endpoint; 7 = right endpoint | Regular |

**Subjective Norm**

Please indicate how much you agree with the following statements.

| **Item** | **Question / bipolar endpoints** | **Scale** | **Coding** |
| --- | --- | --- | --- |
| SN1 | Most people in my environment (e.g., friends, school, university, or work) would approve of me not smoking in the next four weeks. | 1 = strongly disagree; 7 = strongly agree | Regular |
| SN2 | If I am in a situation where others are smoking in the next four weeks, most people in my environment would expect me not to smoke. | 1 = strongly disagree; 7 = strongly agree | Regular |
| SN3 | Most people in my environment would want me to avoid exposure to secondhand smoke in the next four weeks. | 1 = strongly disagree; 7 = strongly agree | Regular |
| SN4* | Most people in my environment would disapprove of me not smoking in the next four weeks. | 1 = strongly disagree; 7 = strongly agree | Reverse-coded |

**Perceived Behavioral Control**

Please indicate how much you agree with the following statements.

| **Item** | **Question / bipolar endpoints** | **Scale** | **Coding** |
| --- | --- | --- | --- |
| PBC1 | Whether I smoke or not in the next four weeks is largely up to me. | 1 = strongly disagree; 7 = strongly agree | Regular |
| PBC2 | Even if I felt tempted to smoke in the next four weeks, I could skip a cigarette/vape or delay smoking. | 1 = strongly disagree; 7 = strongly agree | Regular |
| PBC3 | If someone offers me a cigarette/vape, I can clearly say no in the next four weeks. | 1 = strongly disagree; 7 = strongly agree | Regular |
| PBC4 | In situations with peer pressure (e.g., at a party or during a break), I would be able to stick to not smoking in the next four weeks. | 1 = strongly disagree; 7 = strongly agree | Regular |
| PBC5 | If people in my environment (friends/school/university/work) smoke near me, I can keep my distance or leave in the next four weeks. | 1 = strongly disagree; 7 = strongly agree | Regular |
