## Supplementary figures and images for "Generative AI avatar videos for tobacco prevention on social media: a randomized controlled trial"

### Appendix 2.png

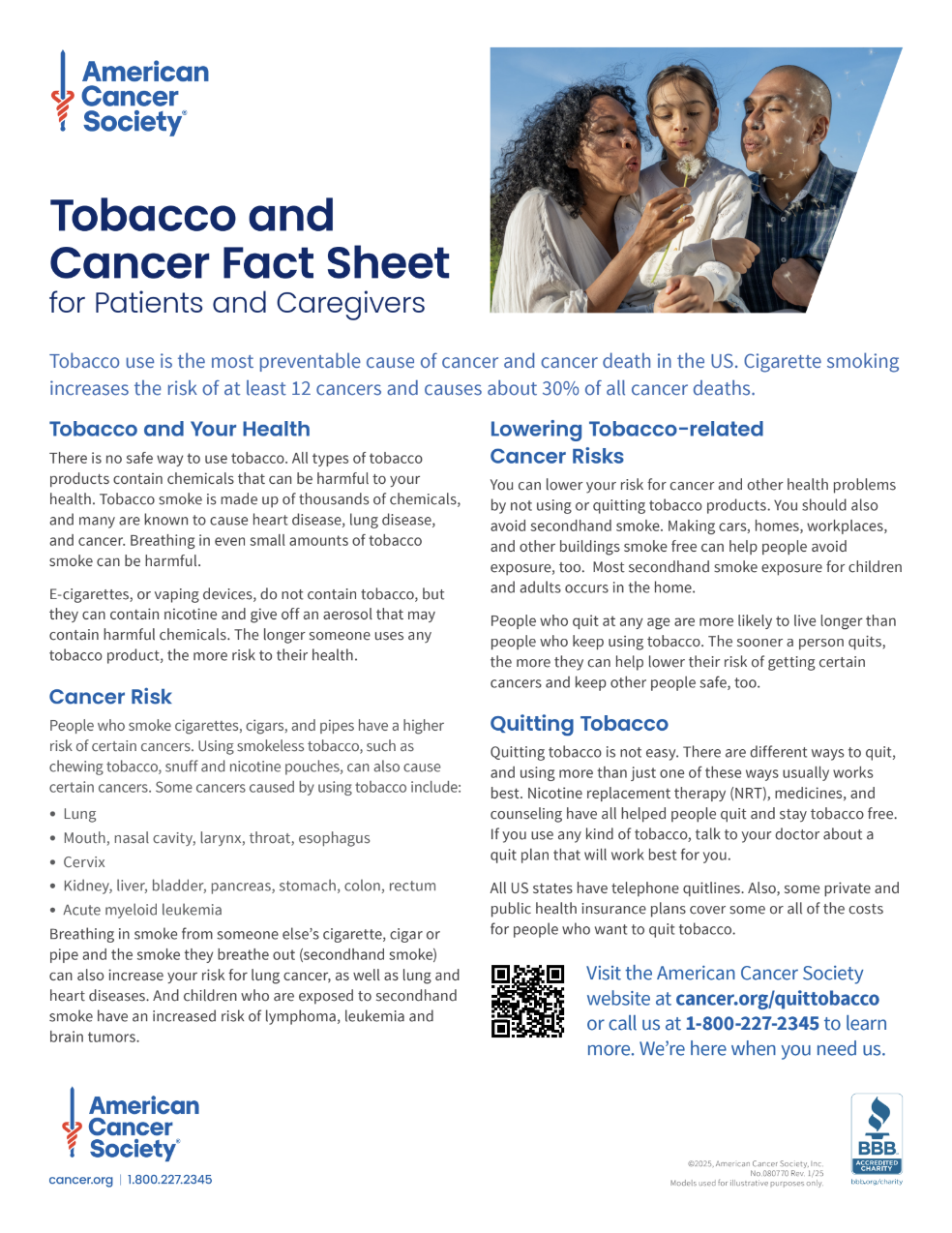
